# Exposure to and engagement with digital psychoeducational content and community related to maternal mental health by perinatal persons and mothers: design of an online survey with optional follow-up and participant characteristics

**DOI:** 10.1101/2024.07.08.24310070

**Authors:** Molly E. Waring, Katherine E. McManus-Shipp, Christiana M. Field, Sandesh Bhusal, Asley Perez, Olivia Shapiro, Sophia A. Gaspard, Cindy-Lee Dennis

## Abstract

**Background:** Leveraging digital platforms may be an effective strategy for connecting perinatal persons and mothers with evidence-based information and support related to maternal mental health and peers. Momwell is a mom-centered model of care that provides psychoeducational content through several digital platforms including social media, podcasts, and blog posts.

**Objective:** To describe the design of a study of perinatal persons and mothers who are exposed to or engage with psychoeducation content and community related to maternal mental health on social media or other digital platforms (Momwell), and to describe characteristics of the sample.

**Methods:** Adults who engaged with Momwell on any of their digital platforms were recruited to participate in an online survey study in summer/fall 2023. Participants completed either a longer or shorter survey. Two to 3 months after completing this survey, participants who provided permission to be re-contacted were invited to complete a second survey. The surveys included validated psychological measures, study-specific quantitative questions, and open-ended questions that assessed participant demographics, exposure to and engagement with Momwell psychoeducation content and community, maternal mental health, parenting relationships, parenting self-efficacy, and additional psychosocial and health measures.

**Results:** Participants (N=584; n=298 longer survey, n=286 shorter survey) were >99% mothers, 46% perinatal (10% pregnant, 36% post-partum), and on average 32.4 (SD: 3.9) years old. Fifty-nine percent were from the United States, 36% from Canada, and 5% from other countries. The vast majority (95%) followed Momwell on Instagram, 44% listened to the Momwell podcast and 41% received their newsletter. Most participants had been exposed to Momwell’s psychoeducation content for at least 6 months across the different platforms (range: 40% TikTok to 87% Instagram). Two to 3 months later, 246 participants completed a second survey (n=149 longer survey, n=97 shorter survey).

**Conclusions:** Data from this study will provide insights into how perinatal persons and mothers leverage digital psychoeducational content and peer communities to support their mental health across the perinatal period and into the early years of motherhood. Leveraging digital platforms to disseminate evidence-based digital psychoeducational content related to maternal mental health and connect peers has the potential to change how we care for perinatal persons and mothers.

## Introduction

Depression and anxiety are common during pregnancy and the post-partum period, yet many struggling with their mental health do not seek mental health care [1,2]. Mothers often feel pressure to achieve socially constructed ideals of motherhood and struggle to balance life roles and responsibilities, which together can negatively impact their mental health [2,3]. Prevention, screening, and treatment of perinatal mental health problems are critical given the long-lasting impact perinatal mental health can have on mothers, children, and families. Several meta-analyses and systematic reviews have shown that psychosocial, psychological, and psychoeducational interventions can improve mood and reduce risk of depression among perinatal persons [4–6] and parents of young children [7]. A recent meta-analysis found that peer support interventions are efficacious for preventing or managing perinatal and post-partum depression [8]. Specifically, information and support from peers via phone can prevent post-partum depression in high-risk women [9], and online asynchronous support groups can also reduce depressive symptoms during the perinatal period [10].

Leveraging digital platforms such as social media and podcasts may be an effective strategy for overcoming barriers to seeking mental health care and connecting perinatal persons and mothers with evidence-based information and peer support related to maternal mental health [11]. The majority of women of childbearing age and mothers use social media, including those in Canada [12] and the United States [13–15], and many turn to their online networks for support and information about diverse health and parenting topics [16,17]. Additionally, 67% of US adults aged 18-39 years old and 58% of those aged 30-49 years listen to podcasts [18]. Connecting with other perinatal persons or mothers online can decrease feelings of isolation, provide a safe space to discuss mental health without feeling judged or stigmatized, and increase parenting confidence [19,20]. Given the ever-expanding opportunities for psychoeducation and peer support available via social media and other digital platforms such as podcasts, content disseminated via these digital platforms has the potential to reach millions, for widespread impact on perinatal and maternal mental health. However, little is known about how perinatal persons and mothers consume and engage with psychoeducational content disseminated through digital platforms, and how this impacts perinatal and maternal mental health. There is an ongoing need to understand how people access and use digital mental health resources focused on perinatal and parenting populations.

The overarching aims of this project were to describe how perinatal persons and mothers engage with psychoeducation content and community, to describe perceived benefits of exposure to and engagement with content and community, to examine associations between engagement with digital psychoeducation content and maternal mental health, parenting attitudes, and interparental relationships, and to examine changes in mental health and parenting attitudes and concurrent engagement in digital psychoeducation content and community over 2-3 months. In this paper, we describe the design of the study and the characteristics of the sample. As the majority of women of childbearing age and mothers in the United States and Canada use social media [12–15] and listen to podcasts [18], there is great potential for wide dissemination of relevant and impactful evidenced-based psychoeducational content that can support maternal mental health.

## Methods

### Overview

We recruited adults exposed to or who engage with Momwell via social media or other digital platforms to complete an online survey. We invited participants who provided permission for us to re-contact them to complete a second survey 2-3 months later. The University of Connecticut Institutional Review Board (IRB) approved this study.

### Momwell

Founded and led by Erica Djossa, a registered psychotherapist based in Ontario, Canada, Momwell offers a mom-centered model of care that seeks to educate, empower, and support mothers (momwell.com). Momwell offers individual telehealth psychotherapy to clients in Canada and the United States and self-paced virtual workshops and courses led by Ms. Djossa and her team of licensed psychotherapists. In addition to these for-fee services, Momwell provides evidence-based psychoeducational content about motherhood, parenting, and maternal mental health to the public without cost through their social media feeds (Instagram, Facebook, and TikTok), weekly podcast, and blog with posts corresponding to the podcast episodes. A weekly email newsletter alerts readers to new podcast episodes and blog posts, and provides other updates including new workshop/course offerings. In the current study, we focused on exposure to and engagement with Momwell psychoeducational content and community via their social media feeds, podcast, blog, and email newsletter (i.e., resources freely available to the public).

### Recruitment and Eligibility Screening

In July-September 2023, we recruited adults who follow Momwell on any of their social media platforms, listen to their podcast, read their blog, and/or receive their email newsletter. The Momwell team posted recruitment messages that included a link to an eligibility screener via Qualtrics. Recruitment messages were posted as public social media posts and Instagram Stories. On Instagram, interested individuals could comment “STUDY” to be sent a link to the eligibility screener via direct message. An ad was included in a Momwell podcast in mid-July 2023. Additionally, a message about the study was included in Momwell’s weekly email newsletter. Individuals were recruited to participate in two cohorts (Cohort 1 and Cohort 2); participants in Cohort 2 completed shorter surveys at both timepoints. Inclusion criteria for both cohorts included age 18 years or older, follows or engages with Momwell on at least one of their digital platforms (e.g., Instagram, Facebook, Facebook group, TikTok, podcast, blog, and/or email newsletters), comfortable participating in the study in English, and able and willing to provide informed consent. To participate in Cohort 1, individuals additionally had to be (1) a perinatal person (e.g., currently pregnant and/or within 12 months post-partum) and/or mother (e.g., identifies as mother of at least one child under the age of 18 who lives with them at least part-time), and (2) currently living in Canada or the United States. As part of the eligibility screener, respondents indicated whether they had a preference of completing the longer or shorter survey (or had no preference); participants who expressed preference for the shorter survey were all invited to complete the shorter survey.

As some recruitment messages were posted publicly, we recognized the potential for attracting the attention of bots or individuals trying to participate under false pretenses in order to receive the gift card incentive [21–24]. As recommended [23,24], we employed a multi-faceted strategy to ensure that enrolled participants were truly eligible, including bot/fraud detection features of Qualtrics, duplicate respondents (e.g., email, IP address), and checking consistency of information (e.g., specific recruitment link clicked on versus where respondent reported they heard about the study). Individuals whose responses to the eligibility screener suggested that they were a bot or fraudulent individual were not invited to complete the survey.

### Data collection

Eligible individuals were emailed invitations to complete an online survey via Qualtrics. Individuals who had not responded to the invitation were sent a reminder one day later. Non-responders were sent two additional invitations a few weeks later. Participants who partially completed the survey were sent a reminder email with a new survey link. Prior to completing the survey, participants were shown an Information Sheet (specific for Cohort 1 and Cohort 2) and provided an opportunity to download a PDF of the Information Sheet. Participants consented to the research study electronically, and were then directed to the survey. The survey included both quantitative and open-ended questions. The survey for Cohort 1 participants was designed to take 30-45 minutes to complete (median [IQR]: 52 [36–95] minutes among those who completed at least 80% of the survey). Cohort 1 participants who completed at least 80% of the survey received a gift card ($15 USD or $20 CAD). The survey for Cohort 2 participants was a shortened version of the Cohort 1 survey, comprised of a subset of questions and scales. It was designed to take 15-20 minutes to complete (median [IQR]: 29 [21–45] minutes among completers). Cohort 2 participants who completed at least 80% of the survey were entered into a gift card lottery. One in every 30 participants were randomly selected to receive a gift card ($60 USD or $80 CAD).

We reviewed participants’ responses and survey meta-data to detect likely fraudulent respondents that were not detected at eligibility screening (e.g., discrepancies between meta-data and survey responses, discrepancies between data provided in eligibility screener and survey). As recommended,[23,24] we conducted additional data quality checks at this phase of data collection; we reviewed surveys for patterns of responses indicating inattentiveness or fraudulent responses (e.g., straight-lined psychological measures with reversed items; failed an attention check item; missing, extremely brief, or nonsensical responses to open-ended questions). We contacted participants with multiple flags via phone and/or email to clarify their responses or ask them to provide responses to the open-ended survey questions. Individuals we could not reach or who provided conflicting responses were excluded.

The final question in the survey asked participants for permission to contact them about future research studies, specifically a second survey in about 2 months. Participants who responded affirmatively were invited to complete an optional second survey 2-3 months later (October-December 2023). Study procedures (e.g., survey invitations, reminders, consent process) were the same as for the main survey. Survey response review was also similar to the main survey; we additionally compared consistency of responses in the two surveys (e.g., pregnancy/post-partum status of participants who were pregnant when they completed the main survey). Similar to the main survey, the second surveys were designed to take 30-45 minutes (Cohort 1; median [IQR] duration: 34 [24–62] minutes among those who completed at least 80% of the survey) and 15-20 minutes (Cohort 2; median [IQR] duration: 12 [9–22] minutes among those who completed at least 80% of the survey). As with the main survey, Cohort 1 participants received a gift card after completing the survey ($15 USD or $20 CAD), and Cohort 2 participants who completed the survey were entered into a gift card lottery in which 1 in 30 participants were randomly s elected to receive a gift card ($60 USD or $80 CAD).

To ensure that participants had opportunity to be exposed to and engage with content related to perinatal mental health, the Momwell content development team created and posted 3 psychoeducational posts specifically related to perinatal mental health per week for 16 weeks starting at the start of recruitment for the main survey.

### Measures

The surveys included a rich set of validated psychological measures, study-specific quantitative questions, and open-ended questions that assessed exposure to and engagement with Momwell psychoeducation content and community, maternal mental health, parenting relationships, parenting self-efficacy, and other participant characteristics and behaviors. Measures are described below and in Table 1.

**Table 1:** Measures assessed, by cohort and time point.

| Measures | Main survey |  | Optional second survey |  |
| --- | --- | --- | --- | --- |
|  | Cohort 1 | Cohort 2 | Cohort 1 | Cohort 2 |
| **Exposure to and engagement with Momwell psychoeducational content and community** |  |  |  |  |
| Exposure to and engagement with Momwell content and community | X | X | X | X |
| Perceived impact of Momwell content | X | X | X | X |
| Suggestions to increase engagement and desired topics for future posts/podcasts | X | X |  |  |
| **Mental health** |  |  |  |  |
| Patient Health Questionnaire (PHQ-8) [25,26] | X | X | X | X |
| General Anxiety Disorder (GAD-7) [27] | X | X | X | X |
| Perceived Stress Scale (PSS-10) [28] | X | X | X | X |
| **Parenting support & self-efficacy** |  |  |  |  |
| Postpartum Partner Support Scale (PPSS) [29] | X |  | X |  |
| Brief Co-Parenting Relationship Scale (BCRS) [30] | X |  | X |  |
| Parenting Sense of Competence Scale (PSOC) [31] | X |  | X |  |
| **Additional psychological measures** |  |  |  |  |
| Ten-Item Personality Inventory (TIPI) [32] | X | X |  |  |
| General Help-Seeking Questionnaire (GHSQ) [33,34] | X |  | X |  |
| Mental Health Literacy Scale (MHLS) [35] | X |  | X |  |
| Brief COPE [36] | X |  | X |  |
| Iowa-Netherlands Comparison Orientation Measure (INCOM)[37–39] | X |  | X |  |
| Self-Compassion Scale Short-Form (SCS-SF) [40,41] | X |  | X |  |
| UCLA Loneliness Scale (ULS-8) [42,43] | X |  | X |  |
| **Additional participant characteristics** |  |  |  |  |
| General social media use and digital health literacy | X | X |  |  |
| Demographics and household composition | X | X | X | X |
| Health and health behaviors | X | X | X | X |
| Health care utilization | X | X | X | X |

### Exposure to and engagement with Momwell psychoeducational content and community

As part of the eligibility screener, main survey, and second survey, participants were asked a variety of questions about their exposure to and engagement with Momwell content and community to capture different dimensions of exposure and engagement including duration, frequency, and depth of active engagement (Table 1).

In the eligibility screener, participants were asked whether they listen to the Momwell podcast, follow Momwell on Instagram, Facebook, or TikTok, are a member of the “Momwell Community” Facebook group, follow the Momwell blog, or receive the Momwell email newsletters. From this we categorized participants as social media followers (i.e., follows Momwell on Instagram, Facebook, or TikTok, or is a member of the Momwell Facebook group) or not. We also categorized whether participants listen to the Momwell podcast and/or read the Momwell blog, as both these platforms provide a deeper dive into psychoeducational topics, and blog posts are published that correspond to each podcast episode.

Those reporting exposure to content on each platform were asked how recently they started consuming content from that platform (within the past 7 days, at least 7 days ago but less than 3 months ago, at least 3 months ago but less than 6 months ago, at least 6 months ago but less than 12 months ago, and at least 12 months ago). As Momwell rebranded themselves as “Momwell” from “Happy as a Mother” in January 2023, we prompted participants to include involvement with “Happy as a Mother”. To capture duration of exposure to Momwell content, we calculated the longest duration of exposure on any platform (i.e., how long ago they joined via their first digital platform). We also calculated how recently they joined a new platform.

For each platform, participants reported how often they engaged in specific ways applicable to that platform within the past 4 weeks (or since they started following Momwell on that platform if more recently than 4 weeks ago) on a 4-item Likert scale (“every day”, “3+ times per week but not every day”, “1-2 times”, and “not at all”). Activities included passive exposure to content (e.g., read posts, watch videos, listen to podcast, read blog) and more active and visible forms of engagement (e.g., liking/reacting to posts, replying to post).

Participants who completed the second survey who did not report exposure to Momwell content on a particular platform at eligibility screening were asked about exposure to Momwell content on that platform during the second survey. Then, existing and new users of each platform were asked the same questions about their exposure to and engagement with Momwell content and community since they completed the main survey, with the recall period of since they completed the first survey (e.g., in [MONTH] 2023, e.g., August 2023).

On the main survey, participants were asked several questions related to their perceived impact of Momwell content on their lives. First, they were asked “what Momwell social media post, blog post, or podcast has had the biggest impact on your life? How did it impact you?”.

On the second survey, they were asked similar open-ended questions about the perceived impact of Momwell content, but specific to each platform. In both surveys, participants were also asked whether they had taken several actions as a result of their exposure to Momwell content (e.g., talked with a doctor or health care professional about something you heard in the Momwell podcast or saw on a social media post). In the main survey, participants reported the extent to which they agreed with several statements about changes since joining the Momwell community (e.g., I feel more aware of the signs and symptoms of mental health conditions, I feel more confident in my approach to parenting). In the main survey, participants were also asked what would help them engage more with the Momwell community and what topics or content they would like to see more of from Momwell.

While our primary interest was in exposure to and engagement with the free psychoeducational content and peer community provided by Momwell through their social media feeds, podcast, and blog, we also asked participants about paid services available from Momwell. Specifically, we asked participants who had joined the Momwell community more than 7 days ago whether they had ever purchased a guided journal or other self-paced tool from Momwell, enrolled in a Momwell workshop or course, or enrolled in therapy (in-person or remote) with a Momwell therapist. From responses, we calculated the proportion who had purchased any paid services.

### Mental health

Our main measures of maternal mental health are depression, anxiety, and perceived stress. Participants in both cohorts were asked to complete these measures in the main and second surveys (Table 1). Depressive symptoms were assessed using the 8-item Patient Health Questionnaire (PHQ-8) [25,26]. The scale asks participants to indicate on a 4-point scale how often they have been bothered by various depressive symptoms in the past 2 weeks. Response options include “Not at all”, “Several days”, “More than half the days”, and “Nearly every day”. Responses are summed to create a total score ranging from 0 to 24 with a greater score indicating greater reported depressive symptoms. Internal consistency was acceptable, with a Cronbach’s α = 0.85 in the main survey and α = 0.86 in the second compared to the original measure (α = 0.86-0.89 across samples) [25].

Symptoms of anxiety were assessed using the Generalized Anxiety Disorder 7-item scale (GAD-7) [27]. Participants were asked to report how often they had been bothered by symptoms of anxiety over the past 2 weeks. Response options include “Not at all”, “Several days”, “More than half the days”, and “Nearly every day”. Responses were summed to create a total score ranging from 0 to 21 with a greater score indicating higher symptoms of anxiety. Internal consistency was acceptable, with a Cronbach’s α = 0.91 in the main survey and α = 0.90 in the second compared to the original measure (α = 0.92) [27]. Participants also reported whether they had ever been diagnosed with depression or anxiety during pregnancy, the post-partum period, or a non-perinatal period of life. Participants reporting a history of depression (or anxiety) were asked if they currently had depression (or anxiety).

Participants who scored 10 or higher on the PHQ8 or GAD7 or who endorsed feeling as if they were currently experiencing depression or anxiety were flagged and shown a message noting that they might be experiencing some feelings of depression (or anxiety, or depression and/or anxiety, depending on which was flagged) and encouraging them to connect with a mental health care professional for assessment, support, and therapy if appropriate, and that their primary care provider or obstetrician/gynecologist may also be a good resource for support or to help you connect with a mental health care professional. Participants were then offered the option of downloading a PDF mental health resource guide developed by the research team (options for participants from the US, Canada, and other countries). Participants were asked if they wanted a copy of the guide sent to them via email, and research staff emailed a copy of the appropriate mental health guide to participants answering affirmatively.

The 10-item Perceived Stress Scale (PSS-10)[28] was used to assess perceived stress during the past 2 weeks. Participants selected how often they experienced the feelings and thoughts presented in the items via response options ranging from “Never” to “Very often”. Responses are summed to create a total score with a range from 0 to 40 with a higher score indicating higher perceived stress. Internal consistency was acceptable, with a Cronbach’s α = 0.88 in the main survey and α = 0.88 in the second compared to the original measure (α = 0.78) [28].

### Parenting support and self-efficacy

Participants in Cohort 1 completed a set of measures related to parenting support and self-efficacy (Table 1). We first asked participants with children whether they had a parenting partner (i.e., a partner who is assisting the participant with parenting, with a note that for many people, their parenting partner is their spouse or the child’s other parent).

Participants who reported a parenting partner completed a modified version of the Postpartum Partner Support Scale (PPSS) [29], a validated 20-item measure that assesses support provided by their husbands or partner. To better represent a variety of partnerships and family structures, we modified the PPSS to replace “him” (i.e., husband) with “my partner”. We also modified questions to refer to “our children” rather than “our baby”. Responses are summed to create a total score with a range from 20 to 80 with higher scores indicating higher parenting support. Internal consistency was acceptable, with a Cronbach’s α = 0.95 in the main survey and α = 0.96 in the second compared to the original measure (α = 0.96) [29].

Participants who reported a parenting partner completed a modified version of the 14-item Brief Co-Parenting Relationship Scale (BCRS) [30] to assess interparental relationships. We modified the phrasing of items and measure instructions to refer to “child(ren)”versus “child” to better reflect families of various sizes. Responses are averaged, with scores ranging from 0-6, with higher scores indicating more positive relationship with parenting partner. Internal consistency was acceptable, with a Cronbach’s α = 0.90 in the main survey and α = 0.91 in the second survey compared to the original measure (α = 0.81-0.89 across different samples) [30].

Perceived parenting competence was assessed using a modified version of the 16-item Parental Sense of Competence (PSOC) scale [31]. We modified the language of the PSOC to replace the words “mother” and “father” in items with “parent” as well as to replace “child” with “children”. The PSOC is composed of two subscales, efficacy and satisfaction, and combined creates an overall score representing parenting self-esteem. The overall PSOC score is reflective of 16-items with a range from 16 to 96 and higher scores indicate greater sense of overall parenting self-esteem. The satisfaction subscale is comprised of 9 items with scores ranging from 9 to 54. Higher scores indicate higher levels of parenting satisfaction. The efficacy subscale is assessed with 7 items, scores ranging from 7 to 42, and higher scores indicate more parenting efficacy. Internal consistency was acceptable for the overall score and both subscales, with a Cronbach’s α = 0.80 for the overall score, α = 0.77 for the satisfaction subscale, and α = 0.76 for the efficacy subscale in the main survey and α = 0.83 for the overall score, α = 0.77 for the satisfaction subscale, and α = 0.83 for the efficacy subscale in the second survey compared to the original measure (α = 0.79 for the overall score, α = 0.76 for satisfaction subscale, and α = 0.76 for efficacy subscale) [31].

### Additional psychological measures

We included several additional psychological measures to provide data for secondary analyses of study data (Table 1). As part of the main survey, participants in both cohorts completed the Ten-Item Personality Inventory (TIPI)[32] which assesses the personality traits extraversion, agreeableness, conscientiousness, emotional stability, and openness to experience. Participants in Cohort 1 completed several additional measures as part of the main and second surveys (Table 1). Participants completed a modified General Help-Seeking Questionnaire (GHSQ) [33,34] which asked individuals to rate how likely they would be to seek help from various sources (e.g., partner or spouse, mental health professional) if they were experiencing sadness, low mood, irritability, and other depressive symptoms. Knowledge of mental health conditions and mental health literacy were assessed using a subset of questions from the Mental Health Literacy Scale (MHLS) [35]. Given the interests of the current study (perinatal mental health versus general mental health/illness), we included items assessing knowledge of generalized anxiety disorder (GAD) and major depressive disorder (MDD), the connection between sleep and mood, information-seeking related to perinatal depression, and general attitudes towards people with perinatal depression (stigma).

Participants completed the Iowa-Netherlands Comparison Orientation Measure (INCOM) [37–39], designed to assess frequency of social comparisons. We modified the measure stem to ask about comparisons with “other mothers” rather than “other people” to assess tendency to make social comparisons to other mothers. Participants completed the short form version of the Self-Compassion Scale (SCS-SF) [40,41], a validated questionnaire measuring self-compassion via 6 subscales. Feelings of loneliness were ascertained using the short-form Revised University of California at Los Angeles (UCLA) Loneliness Scale (ULS-8) [42,43]. Finally, participants completed the Brief COPE [36], which assesses 14 coping strategies: self-distraction, active coping, denial, substance use, use of emotional support, use of instrumental support, behavioral disengagement, venting, positive reframing, planning, humor, acceptance, religion, and self-blame.

### General social media use and digital health literacy

Participants reported their use of social media generally, including whether they have accounts on different platforms (i.e., Instagram, Facebook, TikTok), how often they engage with various platforms in different ways (e.g., post a video), and whether they follow social media accounts related to motherhood or parenting, mental health, or another health topic. Participants were also asked whether they subscribe to or regularly listen to podcasts (other than Momwell). We modified two items from the e-Health Literacy Scale (eHEALS) [44] to assess perceived skill and confidence using social media to access health information: “I have the skills I need to evaluate health information I see on social media.” and “I feel confident in using health information from social media to make health decisions for me or my family.” Response options ranged from “Strongly Disagree (1)” to “Strongly Agree (5)”. As a brief screener of health literacy, participants were asked how confident they are filling out medical forms by themselves [45].

### Demographic characteristics, health behaviors, and health care utilization

Participants reported demographic characteristics, including country and state/province of residence, urbanicity of their city/town of residence, gender, sex assigned at birth, and sexual orientation [46]. Race and ethnicity were assessed via an open-ended question: “How would you describe your racial and ethnic background?” Participants living in the US and Canada were also asked to indicate their racial and ethnic background by choosing all that apply from a standardized list respective to each country. Participants from the US were also asked whether they consider themselves Hispanic or Latina/o (yes or no). Participants also reported whether they self-identified as someone who is a visible minority [47]. Participants also reported education, employment status, family structure, the number of children living in the home at least part-time, and the ages of such children. In the second survey, participants were asked about current pregnancy (including gestational age), if they have given birth to a baby in the past 12 months and if yes, how many months ago they gave birth. Financial strain was assessed by asking participants to identify how difficult it has been for them, in the past 30 days, to pay for usual household expenses (e.g., food, rent, car payments, medical expenses) [48]. Participants reported their current employment status by selecting one of the following options: working full-time, working part-time, working but currently on parental leave, stay-at-home parent or homemaker, unemployed, student, and other. Given the small number of participants who endorsed each of these categories, we collapsed unemployed, student, and other employment status.

Participants completed the Hunger Vital Sign (HVS) [49], a validated, 2-item food insecurity screening tool based on the U.S Household Food Security Survey Module. Participants reported if, in the last 12 months, they had ever worried about food running out due to financial hardship, and if there was ever not enough money to get more food when food did run out. Participants who answered either of the two items as “often true” or “sometimes true” (vs “never”) were considered to be at risk of experiencing food insecurity.[49] Participants who completed the second survey reported employment status, food security, financial strain, and changes to household structure occurring since completing the main survey. To put any changes in mental health in context, as part of the second survey we asked participants to reported if they had experienced various life events since the first survey (e.g., spouse or partner died, got divorced or separated, became pregnant, gave birth or adopted a child). Life events were adapted from the Holmes-Rahe Life Stress Inventory[50]; we added “gave birth or adopted a child”. For participants who reported one or more potentially stressful life event, they were asked to rate how stressful that time was for them on a Likert scale from “not at all stressful” to “extremely stressful”.

Participants were asked questions about their health, health behaviors, and health care utilization. Participants self-reported their height and weight and whether they have any physical or mental health conditions requiring frequent medical visits (and if so, what conditions). Participants answered questions regarding diet, alcohol consumption, tobacco use, physical activity, and sleep. To assess diet quality, participants completed the 8-item Starting The Conversation (STC) dietary screener [51]. Participants reported how many days per week they consumed at least one drink of any alcoholic beverage, number of drinks per day they consumed alcohol, and frequency of binge drinking. Participants also reported ever use and current use of cigarettes and e-cigarettes. Participants reported frequency and duration of moderate or greater intensity physical activity in the past 4 weeks. To assess quality of sleep in the last 4 weeks, participants completed a subset of questions from the Pittsburgh Sleep Quality Index (PSQI) [52] relating to sleep duration and self-rated sleep quality. Participants first reported via free response how many hours of sleep they get at night. Participants were then asked to rate their quality of sleep with possible answer options of “Very bad”, “Fairly bad”, “Fairly good”, and “Very good”. Mothers were also asked how often in the past 4 weeks a child had disrupted their sleep. Participants reported any physical or mental health care services accessed via in-person visits, telehealth visits, phone call, email, or via online patient portal in the past 6 months. In the second survey, participants were asked about new medical diagnoses, health care utilization, and sleep.

### Statistical Analysis

The eligibility screener and surveys were administered in Qualtrics (Qualtrics LLC, Provo, UT). We used Research Electronic Data Capture (REDCap) for participant tracking [53]. Analyses were conducted in SAS 9.4 (SAS Institute, Inc, Cary, NC). We described recruitment and enrollment and then characteristics of participants in our final sample, overall and by study cohort. We compared participant characteristics and exposure to and engagement with Momwell content on different platforms in relation to whether participants completed the second survey using chi-squared tests for categorical variables, Wilcoxon rank sum tests for poor physical and mental health days, and t-tests for other continuous variables.

## Results

We collected data from 584 perinatal persons and mothers (n=298 longer survey, n=286 shorter survey; Figure 1). Of these, 246 completed an optional second survey 2-3 months later (n=149 longer survey, n=97 shorter survey; Figure 1).

**Figure 1:**
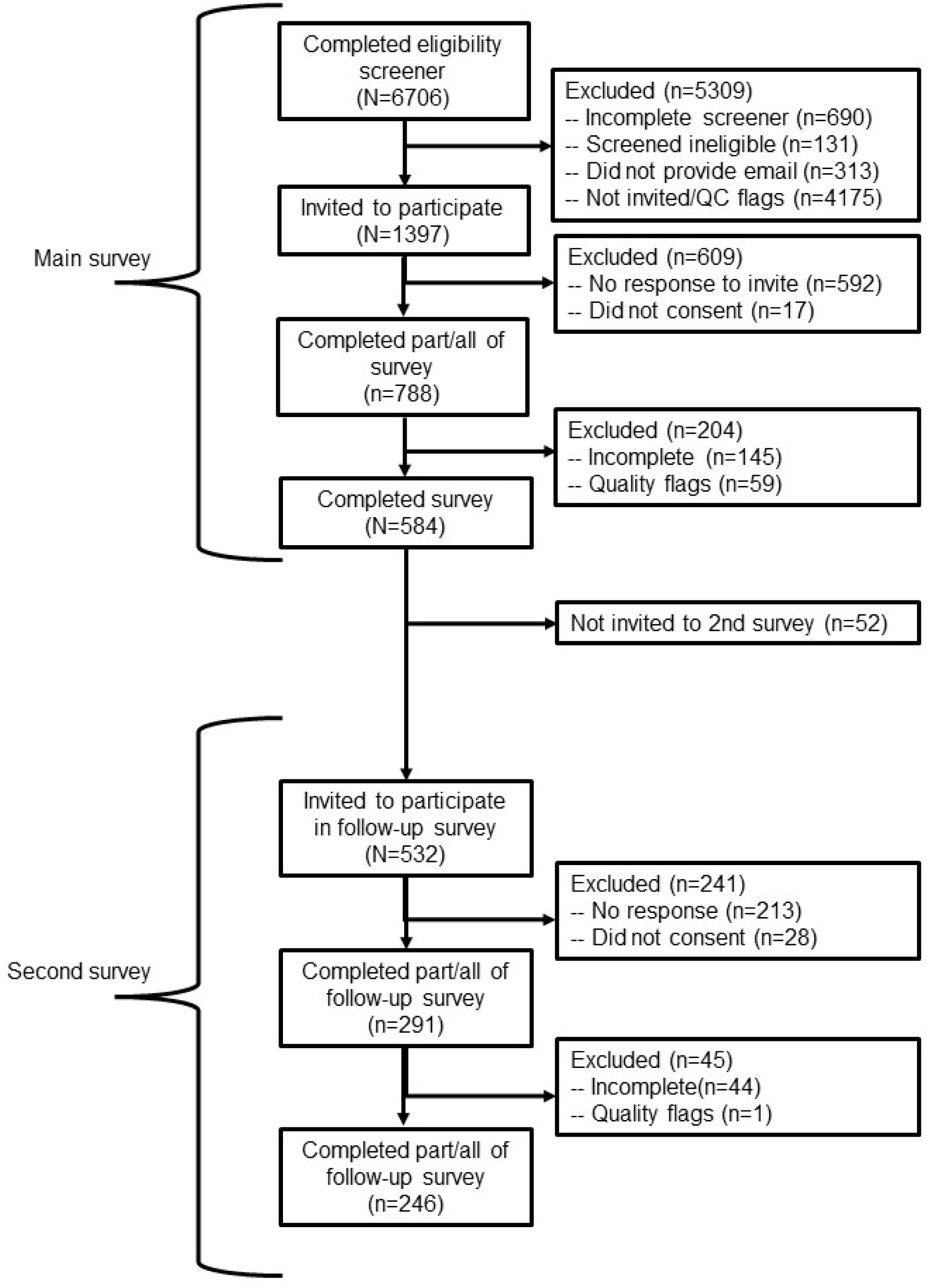
Recruitment and data collection.

One participant in Cohort 2 indicated that they would prefer to not report their gender; all other participants identified as female. Characteristics of the sample, overall and by study cohort, are shown in Table 2. All but 2 participants were mothers (the remaining 2 were pregnant). Almost all participants in Cohort 1 reported that they had a parenting partner (98%; 291 / 296); 97% (283 / 290) of parenting partners were the participant’s spouse or committed partner who lives in same household. Eleven percent of Cohort 2 resided in countries other than the US and Canada, including the UK (n=6), Australia (n=5), Japan (n=2), and 18 other countries (n=1 per country).

**Table 2:** Characteristics of the sample, overall and by study cohort, n (%), M±SD, or median (IQR)

|  | Overall<br>(N=584) | Cohort 1<br>(N=298) | Cohort 2<br>(N=286) |
| --- | --- | --- | --- |
| Identifies as mother | 582 (>99) | 296 (99) | 286 (100) |
| Children 0-5 years | 563 (96) | 285 (96) | 278 (97) |
| Children 6-12 years | 99 (17) | 57 (19) | 42 (15) |
| Children 13-17 years | 22 (4) | 10 (3) | 12 (4) |
| Perinatal status | 266 (46) | 150 (50) | 116 (41) |
| Currently pregnant | 59 (10) | 32 (11) | 27 (9) |
| Currently post-partum | 210 (36) | 120 (40) | 90 (31) |
| Age (years) | 34.2 ± 3.9 | 34.4 ± 3.9 | 34.1 ± 3.8 |
| Country of residence |  |  |  |
| Canada | 208 (36) | 116 (39) | 92 (32) |
| United States | 345 (59) | 182 (61) | 163 (57) |
| Another country | 31 (5) | - | 31 (11) |
| Identifies as a visible minority | 77 (13) | 39 (13) | 38 (13) |
| Urbanicity of residence |  |  |  |
| Large city | 150 (26) | 83 (28) | 67 (24) |
| Suburb near a large city | 212 (36) | 104 (35) | 108 (38) |
| Small city or town | 157 (27) | 74 (25) | 83 (29) |
| Rural area | 63 (11) | 36 (12) | 27 (9) |
| Education |  |  |  |
| Less than Bachelor's | 76 (13) | 38 (13) | 38 (13) |
| Bachelor's (4-year university degree) | 255 (44) | 122 (41) | 133 (47) |
| Graduate degree | 253 (43) | 138 (46) | 115 (40) |
| Employment status |  |  |  |
| Works full-time | 284 (49) | 138 (46) | 146 (51) |
| Works part-time | 91 (16) | 50 (17) | 41 (14) |
| Works but currently on leave | 74 (13) | 40 (13) | 34 (12) |
| Stay-at-home mother | 105 (18) | 54 (18) | 51 (18) |
| Other employment status | 30 (5) | 16 (5) | 14 (5) |
| Difficulty paying for basic expenses |  |  |  |
| Not at all difficult | 333 (57) | 163 (55) | 170 (59) |
| A little difficult | 170 (29) | 86 (29) | 84 (29) |
| Somewhat or very difficult | 80 (14) | 48 (16) | 32 (11) |
| Food insecurity | 63 (11) | 34 (11) | 29 (10) |
| Impaired health literacy | 95 (16) | 48 (16) | 47 (17) |
| Has physical or mental health conditions that require regular medical visits | 151 (26) | 85 (29) | 66 (23) |
| Days in past 4 weeks when physical health was not good | 0 (0-3) | 0 (0-3) | 0 (0-3) |
| Days in past 4 weeks when mental health was not good | 5 (2-10) | 4 (2-8) | 5 (2-10) |
| Social media and podcast use |  |  |  |
| Instagram | 569 (97) | 287 (96) | 282 (99) |
| Facebook | 516 (88) | 266 (89) | 250 (87) |
| TikTok | 139 (24) | 64 (22) | 75 (27) |
| Podcasts | 312 (54) | 162 (55) | 150 (53) |

\* Information missing for some participants (n≤5 per variable)

The majority of the sample (95%, 552 / 584) reported that they follow Momwell on Instagram (Table 3). Roughly 4 in 10 participants listen to the Momwell podcast (44%, 258 / 584) and receive the email newsletter (41%, 240 / 584). Smaller proportions follow Momwell on Facebook or TikTok, are a part of the Momwell Facebook group, or read the blog (Table 3). The largest subgroup – a third (33%, 193 / 584) – followed Momwell on Instagram and did not connect on any other digital platform. Other common combinations on platforms included Instagram and podcast (11%, 66 / 584), Instagram, podcast, and email newsletters (10%, 57 / 584), Instagram and email newsletter (7%, 41 / 584), and Instagram, podcast, blog, and email newsletter (6%, 36 / 584).

**Table 3:**
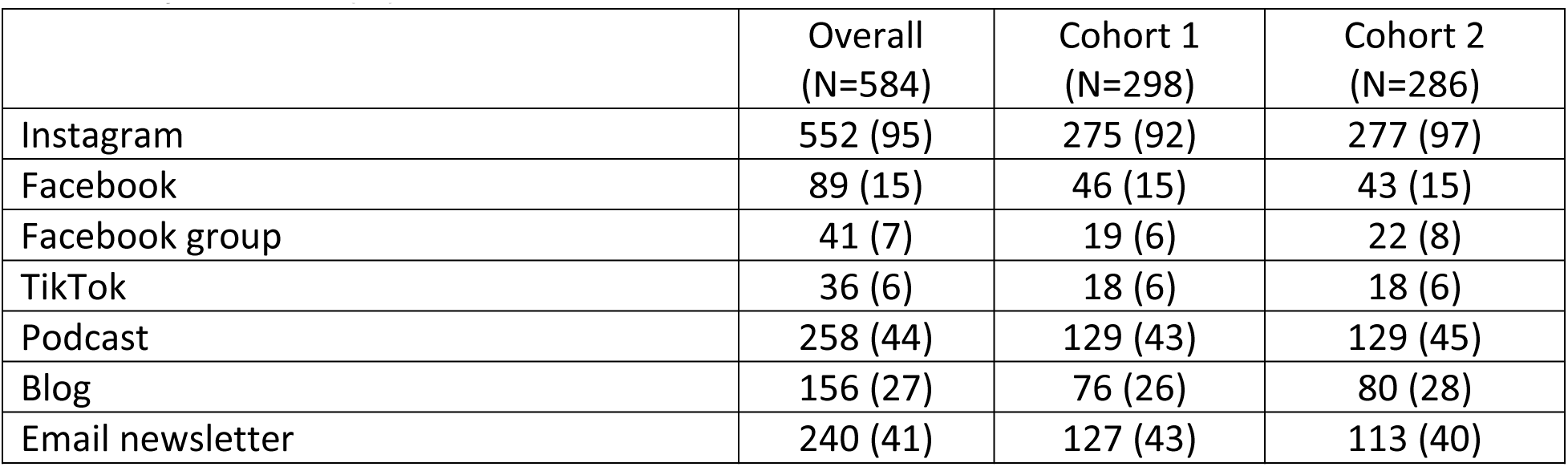
Exposure to Momwell psychoeducational content and community, by digital platform and survey cohort, n (%)

The majority had been exposed to Momwell psychoeducational content and community on each digital platform for at least 6 months: 87% (480 / 552) Instagram, 81% (72 / 89) Facebook, 66% (27 / 41) Facebook group, 44% (16 / 36) TikTok, 60% (154 / 258) podcast, 65% (102 / 156) blog, and 84% (202 / 240) email newsletter.

The majority of the sample (74%, 433 / 584) had been following Momwell on at least 1 digital platform for at least 12 months, and 35% (204 / 584) had started following on a new digital platform within the past 6 months (Table 4). Nearly all participants (97%, 568 / 584) followed Momwell on at least 1 social media platform and 54% (318 / 584) listen to the Momwell podcast and/or read their blog. Thirty percent (175 / 584) were exposed to Momwell content on any platform daily and 52% (306 / 584) were exposed to content 3 or more times per week but not daily (Table 4). Other engagement metrics are summarized in Table 4.

**Table 4:** Exposure to Momwell psychoeducational content and engagement with Momwell community over the past 4 weeks.

| Engagement metric | Overall<br>(N=584) | Cohort 1<br>(N=298) | Cohort 2<br>(N=286) |
| --- | --- | --- | --- |
| Duration of exposure to Momwell content and community |  |  |  |
| Less than 6 months | 61 (10) | 45 (15) | 16 (6) |
| 6+ but less than 12 months | 90 (15) | 50 (17) | 40 (14) |
| At least 12 months | 433 (74) | 203 (68) | 230 (80) |
| How recently started following Momwell on a new digital platform |  |  |  |
| Less than 6 months | 204 (35) | 115 (39) | 89 (31) |
| 6+ but less than 12 months | 121 (21) | 60 (20) | 61 (21) |
| At least 12 months | 259 (44) | 123 (41) | 136 (48) |
| Frequency of exposure to content on any platform |  |  |  |
| Not at all | 3 (<1) | -- | 3 (1) |
| 1-2 times over past 4 weeks | 100 (17) | 56 (19) | 44 (15) |
| 3+ times per week but not daily | 306 (52) | 161 (54) | 145 (51) |
| Daily | 175 (30) | 81 (27) | 94 (33) |
| How often read posts or watched videos on social media in past 4 weeks<sup>a</sup> |  |  |  |
| Not at all | 6 (1) | 1 (<1) | 5 (2) |
| 1-2 times | 94 (17) | 50 (17) | 44 (16) |
| 3+ times per week but not daily | 297 (52) | 157 (55) | 140 (50) |
| Daily | 171 (30) | 78 (27) | 93 (33) |
| How often liked/reacted to social media posts/videos in the past 4 weeks<sup>a</sup> |  |  |  |
| Not at all | 94 (17) | 45 (16) | 49 (17) |
| 1-2 times | 252 (44) | 130 (45) | 122 (43) |
| 3+ times per week | 222 (39) | 111 (39) | 111 (39) |
| Replied to social media posts/videos in the past 4 weeks<sup>a</sup> | 199 (35) | 97 (34) | 102 (36) |
| Listens to podcast or reads blog | 318 (54) | 161 (54) | 157 (55) |
| How often listened to podcast or read blog in past 4 weeks<sup>b</sup> |  |  |  |
| Not at all | 49 (16) | 28 (17) | 21 (14) |
| 1-2 times | 216 (69) | 108 (68) | 108 (70) |
| 3+ times per week | 50 (16) | 24 (15) | 26 (17) |

<sup>a</sup> Among participants who follow Momwell on 1+ social media platform (n=568).
<sup>b</sup> Among participants who listen to the Momwell podcast and/or read the Momwell blog (n=315 / 318; n=3 missing information about listen/read frequency)

A quarter of participants had obtained paid services from Momwell (n=140 / 563, among those who had joined the Momwell community more than 7 days ago); 6% (n=34 / 564 had purchased a guided journal or other self-paced tool from Momwell, 16% (n=92 / 566) had enrolled in a Momwell workshop or course, and 10% (n=59 / 567) had enrolled in therapy with a Momwell therapist.

Main measures of mental health, parenting support, and parenting self-esteem, overall and by study cohort, are shown in Table 5. Overall, 18% (n=105 / 576) of participants had elevated depressive symptoms (n=55 / 293, 19% of cohort 1, n=50 / 283, 18% of cohort 2) and 24% (n=140 / 573) had elevated anxiety symptoms (n=70 / 293, 24% of cohort 1, n=70 / 280, 25% of cohort 2).

**Table 5:**
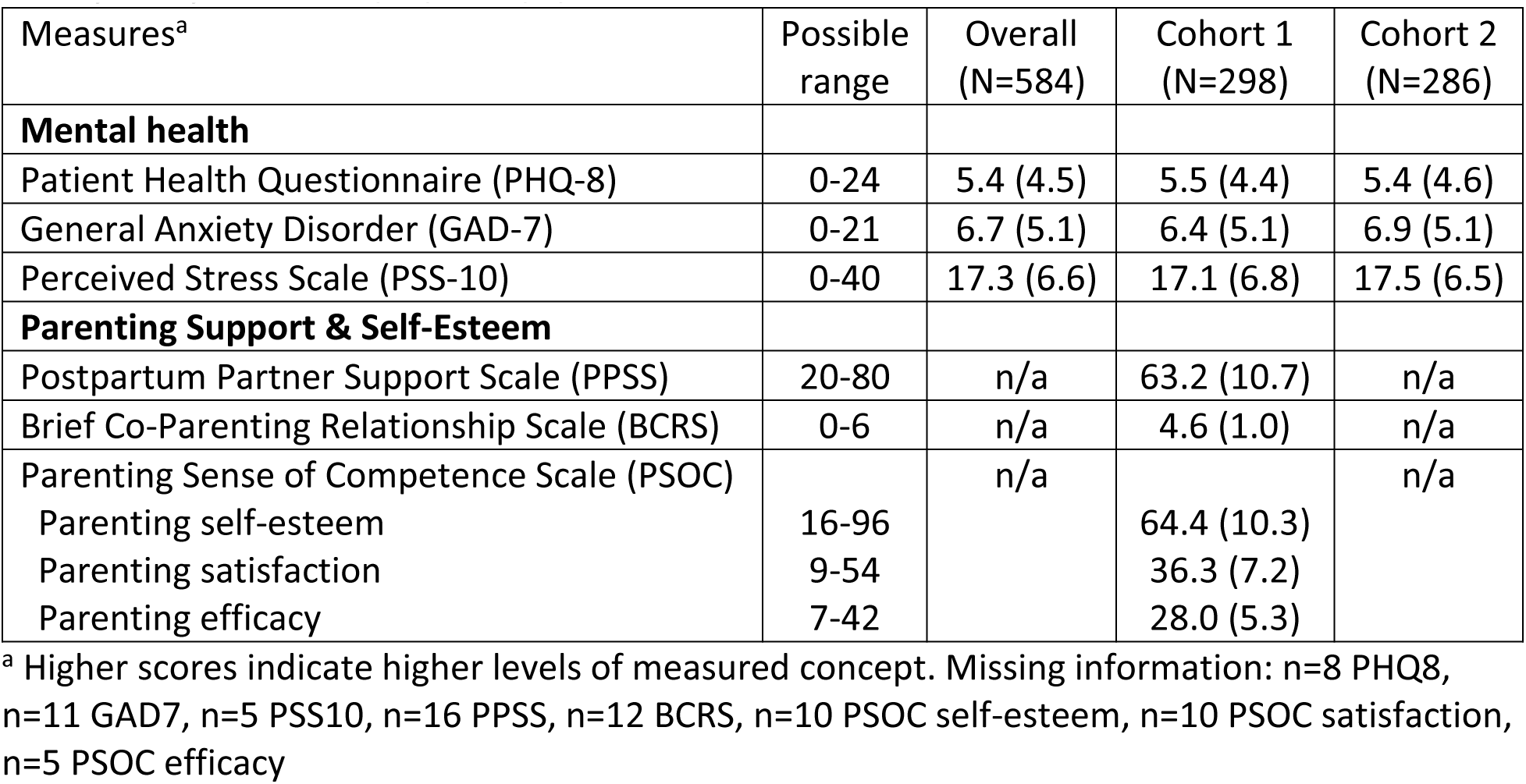
Main mental health, parenting support, and parenting self-efficacy measures, overall and by study cohort, M(SD) or n (%)

Participant characteristics, exposure to and engagement with Momwell content on different digital platforms, mental health, parent support, and parenting efficacy were largely similar among participants who completed just the main survey compared to those who also completed the second survey (all p’s >0.05). Participants who completed the second survey were more likely to have chronic health conditions (30%, n=75 / 246 vs 23%, n=76 / 336, p=0.0324) and more likely to read the Momwell blog (33%, n=82 / 246 vs 22%, n=74 / 338, p=0.0020) than participants who did not complete the second survey. Country of residence also differed by completion of the optional second survey (41%, n=102 / 246 Canada, 55%, n=136 / 246 US, 3%, n=8 / 246 other country among those who completed the second survey vs 31%, n=106 / 338 Canada, 62%, n=209 / 338 US, 7%, n=23 / 338 other country among those who did not complete the second survey, p=0.0143).

## Discussion

### Principal Results

The overall goal of this study is to understand how mothers and the perinatal population consume and engage with digital psychoeducation content and the peer community and how it impacts their mental health and parenting. In this paper, we described the study design including recruitment procedures, characteristics of the sample, and their exposure to maternal mental health psychoeducational content on different digital platforms.

While we recruited a cohort of nearly 600 perinatal persons and mothers, we encountered challenges during recruitment, particularly related to fraudulent and low-quality responses. Bots and fraudulent respondents are becoming more and more of a problem when recruiting participants online, particularly when using public recruitment links and recruiting for studies that provide participant with a monetary incentive for completing an online survey [21–24]. As some of our recruitment messages were posted publicly, we recognized the potential for attracting the attention of bots or individuals trying to participate under false pretenses – and indeed, we experienced significant interest in our study by suspicious actors (human or bot), with several hundred respondents with duplicate email addresses, more than 1,800 with nonsense email addresses, and over 2,000 with conflicting information in terms of location, digital platform engagement, or recruitment source. Bot and fraud detection features of survey administration platforms such as Qualtrics and REDCap (Research Electronic Data Capture) provide a starting point, but researchers must go further to verify the eligibility and quality of respondents. As recommended [23,24], we employed a multi-faceted, multi-stage strategy. We separated eligibility screening from the survey, and only sent individual survey invitations to individuals who passed our reviews. We compared information collected by multiple methods (e.g., recruitment source reported by participant vs recorded by Qualtrics) and consistency of information provided by participants (e.g., ages of children). Our strategy for identifying low-quality or inattentive respondents included an attention check question, reviewing responses to open-ended survey questions, and checking for straight-lining when completing psychological measures [23,24]. We also phrased eligibility questions to reduce the chance that respondents could guess the “right” (i.e., eligible) response. We followed up with participants with inconsistent information on the survey via phone or email to clarify information. While it is possible that we excluded some eligible individuals, our quality control procedures were designed to prevent bots or fraudulent participants from participating in our study. Researchers who are recruiting samples online using public survey links are encouraged to develop and implement similar procedures for preventing and detecting bot and fraudulent respondents. For additional discussion and specific recommendations, researchers are encouraged to read Walker (2023) and Wang (2023), especially their Table 1 and Tables 2 and 4, respectively [23,24].

Another recruitment challenge related to which members of the Momwell community volunteered to participate in this study. We had originally intended to invite only new followers/subscribers (i.e., started following Momwell in the past 7 days) to complete the optional second survey to explore engagement and concurrent changes in mental health and parenting self-efficacy soon after connecting with Momwell content and community. However, only 42 eligible respondents reported joining Momwell within the past 7 days (of whom 15 completed the main survey) – perhaps a result of algorithmic biases in which followers were likely to see our recruitment messages [54,55], and which followers were interested in sharing their experiences with us. Therefore, we pivoted our design to invite any participants to complete the second survey.

Our sample was more likely to have Bachelor’s degree or higher education (87%) than birthing persons in the United States (36%) [56] and women aged 25-34 years born in Canada (40%) [57]. Thirteen percent of our sample identified as a visible minority, compared to 31% of women 25-64 years in Canada [58]; 48% of birthing persons in the United States identify as a race or ethnicity other than non-Hispanic white [56]. In our sample, food insecurity was reported by 11%, which is similar to food insecurity among households with children and married couples in the United States (11%) [59], but lower than the proportion among women aged 25-44 years in Canada (20%) [60]. Whether this is a function of the demographics of the Momwell community or due to which Momwell followers volunteered to participate in this study is unknown.

In the current study, 18% of participants had elevated depressive symptoms and 24% had elevated anxiety symptoms based on their scores on the PHQ8 and GAD7, respectively. These prevalences are similar to estimated prevalences globally and specifically among perinatal persons and mothers in Canada and the United States. The prevalence of depression varies across populations and whether measured by self-report measures of symptoms (as in the current study) or clinical interviews, but estimates are in the 13-28% for perinatal or post-partum persons worldwide [61–63]. Another systematic review and meta-analysis found a prevalence of depression of 16% among healthy mothers without a previous diagnosis of depression from North America [64]. Meta-analyses have estimated the global prevalence of perinatal anxiety to be 21% [65] or 15% during pregnancy and 15% during the first 6 months post-partum [66]. These authors also noted variability across populations sampled and measure of anxiety [65]. The findings of these studies – along with the findings of the current study – that between 1 in 5 perinatal persons or mothers of young children are experiencing elevated symptoms of depression or anxiety highlights the importance of this area of research.

## Limitations

This study has additional limitations worth noting. First, our sample is composed of perinatal persons and mothers who volunteered to participate in this study, and thus likely do not represent all individuals who consume Momwell content. Specifically, more engaged followers (e.g., regularly comment on or like Instagram posts) are more likely to have seen our recruitment messages in their social media feeds [54,55], and those who regularly read the weekly email newsletters or listen to the podcast are more likely to see our recruitment messages disseminated through these communication channels. Followers or community members who feel more connected to the community or perceive higher benefit may be more likely to volunteer to share their experiences, and thus perceived impacts may overrepresent community members who perceive greater positive impact on their lives and wellbeing. Another limitation is the proportion of participants who chose to complete the optional second survey (50% of those who completed the longer survey and 34% of those who completed the shorter survey). The lower response rate for participants who completed the shorter survey may be in part due to the incentive structure, as these participants were entered into a lottery with 1 in 30 selected to receive a gift card. However, demographic characteristics, exposure to Momwell content on different digital platforms, mental health, parent support, and parenting efficacy were overall quite similar among participants who completed just the main survey compared to those who also completed the second survey. Third, the response options for many of the survey questions assessing frequency of engaging with Momwell were “not at all”, “1-2 times”, “3+ times per week but not every day”, and “every day”. After data collection, we realized that these response options were not exhaustive, and participants whose frequency of exposure was between “1-2 times” and “3+ times per week but not every day” over the recall period were forced to pick the more apt of these two options.

## Conclusions

In this paper we describe the design and methods of this study and characteristics of perinatal persons and mothers exposed to or who engage with Momwell content and community. In future papers we will describe perinatal persons’ and mothers’ perceived impacts of this content and peer interactions, associations between exposure to and engagement with digital psychoeducation content and maternal mental health, parenting attitudes, and interparental relationships, and concurrent engagement over 2-3 months and changes in mental health and parenting attitudes. Beyond these planned analyses, we hope that these data can provide additional insights into the health and wellbeing of perinatal persons and mothers. We have shared a de-identified public use dataset with a subset of data collected (variables omitted to protect participant confidentiality) [67], and researchers interested in collaborating with our research team are encouraged to email the first author.

Maternal mental health is critically important, and understanding how individuals, clinicians, and health systems can leverage digital platforms to disseminate evidence-based digital psychoeducational content and connect perinatal persons and mothers with mental health care professionals and peers has potential to change how we care for individuals during these life phases. As the majority of women of childbearing age and mothers in the United States and Canada use social media [12–15] and listen to podcasts [18], there is great potential for wide dissemination of relevant and impactful evidence-based psychoeducational content that can support maternal mental health. We have designed this study to provide insights into how perinatal persons and mothers leverage digital psychoeducational content and communities to support their mental health during pregnancy, the post-partum period, and the early years of motherhood.

## Data Availability

Most of the data presented in this paper are available online at OpenICPSR (https://doi.org/10.3886/E204321V3). All data presented in this paper are available upon reasonable request to the first author.

## Acknowledgements

This study was funded by a grant from the Daymark Foundation (PI: Waring). Vani Jain of the Daymark Foundation and Erica Djossa of Momwell provided input into the design of this study, particularly by providing input into the research questions and concepts to be measured, and Erica Djossa and Momwell staff helped to develop recruitment messages (which were then approved by the University of Connecticut IRB) and disseminated these messages to their followers/subscribers. The Daymark Foundation and Momwell had no additional role in data collection or results reporting.

## Conflicts of Interest

None.

## Abbreviations

BCRS: Brief Co-Parenting Relationship Scale
CAD: Canadian dollar
COPE: Coping Orientation to Problems Experienced
e-HEALS: e-Health Literacy Scale
GAD-7: General Anxiety Disorder
GHSQ: General Help-Seeking Questionnaire
HVS: Hunger Vital Sign
INCOM: Iowa-Netherlands Comparison Orientation Measure
IP: Internet Protocol
IQR: Interquartile Range
MDD: Major Depressive Disorder
MHLS: Mental Health Literacy Scale
PHQ-8: Patient Health Questionnaire
PPSS: Postpartum Partner Support Scale
PSOC: Parenting Sense of Competence Scale
PSQI: Pittsburgh Sleep Quality Index
PSS-10: Perceived Stress Scale
SCS-SF: Self-Compassion Scale-Short Form
SD: Standard Deviation
SDT: Self-Determination Theory
STC: Starting The Conversation
TIPI: Ten-Item Personality Inventory
ULS-8: UCLA Loneliness Scale
US: United States
USD: United States dollar

## Notes

### Competing Interest Statement

The authors have declared no competing interest.

### Author Declarations

The Institutional Review Board (IRB) of the University of Connecticut gave ethical approval for this work.

## References

1. Hahn-Holbrook J, Cornwell-Hinrichs T, Anaya I. Economic and Health Predictors of National Postpartum Depression Prevalence: A Systematic Review, Meta-analysis, and Meta-Regression of 291 Studies from 56 Countries. Front Psychiatry. 2017;8:248.

2. Meeussen L, Van Laar C. Feeling Pressure to Be a Perfect Mother Relates to Parental Burnout and Career Ambitions. Front Psychol. 2018;9:2113.

3. Daehn D, Rudolf S, Pawils S, Renneberg B. Perinatal mental health literacy: knowledge, attitudes, and help-seeking among perinatal women and the public - a systematic review. BMC Pregnancy Childbirth. 2022 Jul 19;22(1):574.

4. Dennis CL, Dowswell T. Psychosocial and psychological interventions for preventing postpartum depression. Cochrane Database Syst Rev. 2013 Feb 28;(2):Cd001134.

5. Park S, Kim J, Oh J, Ahn S. Effects of psychoeducation on the mental health and relationships of pregnant couples: A systemic review and meta-analysis. Int J Nurs Stud. 2020 Apr;104:103439.

6. Branquinho M, Rodriguez-Muñoz MF, Maia BR, Marques M, Matos M, Osma J, et al. Effectiveness of psychological interventions in the treatment of perinatal depression: A systematic review of systematic reviews and meta-analyses. J Affect Disord. 2021 Aug 1;291:294–306.

7. MacKinnon AL, Silang K, Penner K, Zalewski M, Tomfohr-Madsen L, Roos LE. Promoting Mental Health in Parents of Young Children Using eHealth Interventions: A Systematic Review and Meta-analysis. Clin Child Fam Psychol Rev. 2022 Sep;25(3):413–34.

8. Fang Q, Lin L, Chen Q, Yuan Y, Wang S, Zhang Y, et al. Effect of peer support intervention on perinatal depression: A meta-analysis. Gen Hosp Psychiatry. 2022 Feb;74:78–87.

9. Dennis CL, Hodnett E, Kenton L, Weston J, Zupancic J, Stewart DE, et al. Effect of peer support on prevention of postnatal depression among high risk women: multisite randomised controlled trial. BMJ. 2009 Jan 15;338:a3064.

10. Vigod SN, Slyfield Cook G, Macdonald K, Hussain-Shamsy N, Brown HK, de Oliveira C, et al. Mother Matters: Pilot randomized wait-list controlled trial of an online therapist-facilitated discussion board and support group for postpartum depression symptoms. Depress Anxiety. 2021 Aug;38(8):816–25.

11. Fonseca A, Monteiro F, Alves S, Gorayeb R, Canavarro MC. Be a Mom, a Web-Based Intervention to Prevent Postpartum Depression: The Enhancement of Self-Regulatory Skills and Its Association With Postpartum Depressive Symptoms. Front Psychol. 2019;10:265.

12. Schimmele C, Fonberg J, Schellenberg G. Canadians’ assessments of social media in their lives. Stat Can Econ Soc Rep. 2021;1(3).

13. Duggan M, Lenhart A, Lampe C, Ellison NB. Parents and Social Media [Internet]. 2015. Available from: https://www.pewresearch.org/internet/2015/07/16/parents-and-social-media/

14. Waring ME, Blackman Carr LT, Heersping GE. Social Media Use Among Parents and Women of Childbearing Age in the US. Prev Chronic Dis. 2023 Feb 16;20:E07.

15. Gottfried J. Americans’ Social Media Use [Internet]. Pew Research Center; 2024 Jan. Available from: https://www.pewresearch.org/internet/2024/01/31/americans-social-media-use/

16. Ryan R, Davis-Kean P, Bode L, Krüger J, Mneimneh Z, Singh L. Parenting online: analyzing information provided by parenting-focused Twitter accounts. Atl J Commun. 2022;1–17.

17. Adamski M, Truby H, M. Klassen K, Cowan S, Gibson S. Using the Internet: Nutrition Information-Seeking Behaviours of Lay People Enrolled in a Massive Online Nutrition Course. Nutrients. 2020;12(3).

18. Shearer E, Liedke J, Matsa KE, Lipka M, Jurkowitz M. Podcasts as a source of news and information [Internet]. 2023. Available from: https://www.pewresearch.org/journalism/2023/04/18/podcasts-as-a-source-of-news-and-information/

19. Moore D, Drey N, Ayers S. A meta-synthesis of women’s experiences of online forums for maternal mental illness and stigma. Arch Womens Ment Health. 2020 Aug;23(4):507–15.

20. Harrison V, Moore D, Lazard L. Supporting perinatal anxiety in the digital age; a qualitative exploration of stressors and support strategies. BMC Pregnancy Childbirth. 2020 Jun 17;20(1):363.

21. Levi R, Ridberg R, Akers M, Seligman H. Survey Fraud and the Integrity of Web-Based Survey Research. Am J Health Promot AJHP. 2022 Jan;36(1):18–20.

22. Ridge D, Bullock L, Causer H, Fisher T, Hider S, Kingstone T, et al. “Imposter participants” in online qualitative research, a new and increasing threat to data integrity? Health Expect Int J Public Particip Health Care Health Policy. 2023 Jun;26(3):941–4.

23. Walker LO, Murry N, Longoria KD. Improving Data Integrity and Quality From Online Health Surveys of Women With Infant Children. Nurs Res. 2023 Oct 1;72(5):386–91.

24. Wang J, Calderon G, Hager ER, Edwards LV, Berry AA, Liu Y, et al. Identifying and preventing fraudulent responses in online public health surveys: Lessons learned during the COVID-19 pandemic. PLOS Glob Public Health. 2023;3(8):e0001452.

25. Kroenke K, Spitzer RL, Williams JB. The PHQ-9: validity of a brief depression severity measure. J Gen Intern Med. 2001 Sep;16(9):606–13.

26. Kroenke K, Strine TW, Spitzer RL, Williams JB, Berry JT, Mokdad AH. The PHQ-8 as a measure of current depression in the general population. J Affect Disord. 2009 Apr;114(1– 3):163–73.

27. Spitzer RL, Kroenke K, Williams JB, Lowe B. A brief measure for assessing generalized anxiety disorder: the GAD-7. Arch Intern Med. 2006 May 22;166(10):1092–7.

28. Cohen S, Williamson GM. Chapter 3: Perceived Stress in a Probability Sample of the United States. In: Spacapan S, Oskamp S, editors. The Social Psychology of Health. Newbury Park, CA: Sage; 1988. p. 31–67.

29. Dennis CL, Brown HK, Brennenstuhl S. The Postpartum Partner Support Scale: Development, psychometric assessment, and predictive validity in a Canadian prospective cohort. Midwifery. 2017 Nov;54:18–24.

30. Feinberg ME, Brown LD, Kan ML. A Multi-Domain Self-Report Measure of Coparenting. Parent Sci Pr. 2012 Jan 1;12(1):1–21.

31. Johnston C, Mash EJ. A measure of parenting satisfaction and efficacy. J Clin Child Psychol. 1989;18:167–75.

32. Gosling SD, Rentfrow PJ, Swann WB. A very brief measure of the Big-Five personality domains. J Res Personal. 2003 Dec 1;37(6):504–28.

33. Wilson CJ, Deane FP, Ciarrochi J, Rickwood D. Measuring Help-Seeking Intentions: Properties of the General Help-Seeking Questionnaire. Can J Couns. 2005 Jan;39(1):15–28.

34. Rickwood D, Deane FP, Wilson CJ, Ciarrochi J. Young people’s help-seeking for mental health problems. Aust E-J Adv Ment Health. 2005;4(3):218–51.

35. O’Connor M, Casey L. The Mental Health Literacy Scale (MHLS): A new scale-based measure of mental health literacy. Psychiatry Res. 2015 Sep 30;229(1–2):511–6.

36. Carver CS. You want to measure coping but your protocol’s too long: consider the brief COPE. Int J Behav Med. 1997;4(1):92–100.

37. Gibbons FX, Buunk BP. Individual differences in social comparison: development of a scale of social comparison orientation. J Soc Psychol. 1999 Jan;76(1):129–42.

38. Rose JP, Edmonds KA, Gallinari E, Herzog NK, Kumar M. Tendencies for Comparing Up and Down: An Examination of the Directional Subscales of the Iowa-Netherlands Comparison Orientation Measure. J Pers Assess. 2024;106(1):127–43.

39. Butzer B, Kuiper NA. Relationships between the frequency of social comparisons and self-concept clarity, intolerance of uncertainty, anxiety, and depression. Personal Individ Differ. 2006 Jul 1;41(1):167–76.

40. Neff KD. The Development and Validation of a Scale to Measure Self-Compassion. Self Identity. 2003;2(3):223–50.

41. Raes F, Pommier E, Neff KD, Van Gucht D. Construction and factorial validation of a short form of the Self-Compassion Scale. Clin Psychol Amp Psychother. 2011;18(3):250–5.

42. Russell D, Peplau LA, Ferguson ML. Developing a measure of loneliness. J Assess. 1978 Jun;42(3):290–4.

43. Hays RD, DiMatteo MR. A short-form measure of loneliness. J Assess. 1987 Spring;51(1):69–81.

44. Norman CD, Skinner HA. eHEALS: The eHealth Literacy Scale. J Med Internet Res. 2006 Nov 14;8(4):e27.

45. Powers BJ, Trinh JV, Bosworth HB. Can this patient read and understand written health information? Jama. 2010 Jul 7;304(1):76–84.

46. National Academies of Sciences, Engineering, and Medicine. Measuring Sex, Gender Identity, and Sexual Orientation [Internet]. Becker T, Chin M, Bates N, editors. Washington (DC): National Academies Press (US); 2022 [cited 2024 Feb 1]. Available from: http://www.ncbi.nlm.nih.gov/books/NBK578625/

47. Dennis CL, Marini F, Prioreschi A, Dol J, Birken C, Bell RC. The Canadian Healthy Life Trajectories Initiative (HeLTI) Trial: a study protocol for monitoring fidelity of a preconception-lifestyle behaviour intervention. Trials. 2023 Apr 7;24(1):262.

48. Hall MH, Matthews KA, Kravitz HM, Gold EB, Buysse DJ, Bromberger JT, et al. Race and financial strain are independent correlates of sleep in midlife women: the SWAN sleep study. Sleep. 2009 Jan;32(1):73–82.

49. Hager ER, Quigg AM, Black MM, Coleman SM, Heeren T, Rose-Jacobs R, et al. Development and validity of a 2-item screen to identify families at risk for food insecurity. Pediatrics. 2010 Jul;126(1):e26–32.

50. Holmes TH, Rahe RH. The social readjustment rating scale. J Psychosom Res. 1967 Aug 1;11(2):213–8.

51. Paxton AE, Strycker LA, Toobert DJ, Ammerman AS, Glasgow RE. Starting the conversation performance of a brief dietary assessment and intervention tool for health professionals. Am J Prev Med. 2011 Jan;40(1):67–71.

52. Buysse DJ, Reynolds CF, Monk TH, Berman SR, Kupfer DJ. The Pittsburgh Sleep Quality Index: a new instrument for psychiatric practice and research. Psychiatry Res. 1989;28(2):193–213.

53. Harris PA, Taylor R, Thielke R, Payne J, Gonzalez N, Conde JG. Research electronic data capture (REDCap)--a metadata-driven methodology and workflow process for providing translational research informatics support. J Biomed Inf. 2009 Apr;42(2):377–81.

54. Mosseri A. Instagram Ranking Explained | Instagram Blog [Internet]. 2023 [cited 2024 Mar 28]. Available from: https://about.instagram.com/blog/announcements/instagram-ranking-explained

55. Meta. Our Approach to Facebook Feed Ranking [Internet]. 2023 [cited 2024 Mar 28]. Available from: https://transparency.fb.com/features/ranking-and-content/

56. Osterman MJK, Hamilton BE, Martin JA, Driscoll AK, Valenzuela CP. Births: Final Data for 2021. Natl Vital Stat Rep Cent Dis Control Prev Natl Cent Health Stat Natl Vital Stat Syst. 2023 Jan;72(1):1–53.

57. Statistics Canada. Canada leads the G7 for the most educated workforce, thanks to immigrants, young adults and a strong college sector, but is experiencing significant losses in apprenticeship certificate holders in key trades [Internet]. 2022 [cited 2024 May 15]. Available from: https://www150.statcan.gc.ca/n1/en/daily-quotidien/221130/dq221130a-eng.pdf

58. Statistics Canada. Special Interest Profile, 2021, Census of Population, Profile of interest: Visible minority [Internet]. 2024. Available from: https://www12.statcan.gc.ca/census-recensement/2021/dp-pd/sip/details/page.cfm?GenerationStatusId=1&Dguid=2021A000011124&AgeId=6&Lang=E&FocusId=1&PoiId=9&TId=12#sipTable

59. Rabbitt MP, Hales LJ, Burke MP, Coleman-Jensen A. Statistical Supplement to Household Food Security in the United States in 2022 [Internet]. U.S. Department of Agriculture, Economic Research Service; 2023. Report No.: AP-119. Available from: https://www.ers.usda.gov/webdocs/publications/107710/ap-119.pdf?v=1282.9

60. Statistics Canada. Table 13-10-0835-01 Food insecurity by selected demographic characteristics [Internet]. 2024 [cited 2024 May 15]. Available from: https://www150.statcan.gc.ca/t1/tbl1/en/tv.action?pid=1310083501

61. Bauman BL, Ko JY, Cox S, D’Angelo Mph DV, Warner L, Folger S, et al. Vital Signs: Postpartum Depressive Symptoms and Provider Discussions About Perinatal Depression - United States, 2018. MMWR Morb Mortal Wkly Rep. 2020 May 15;69(19):575–81.

62. Al-Abri K, Edge D, Armitage CJ. Prevalence and correlates of perinatal depression. Soc Psychiatry Psychiatr Epidemiol. 2023 Nov;58(11):1581–90.

63. Fairbrother N, Janssen P, Antony MM, Tucker E, Young AH. Perinatal anxiety disorder prevalence and incidence. J Affect Disord. 2016 Aug;200:148–55.

64. Shorey S, Chee CYI, Ng ED, Chan YH, Tam WWS, Chong YS. Prevalence and incidence of postpartum depression among healthy mothers: A systematic review and meta-analysis. J Psychiatr Res. 2018 Sep;104:235–48.

65. Fawcett EJ, Fairbrother N, Cox ML, White IR, Fawcett JM. The Prevalence of Anxiety Disorders During Pregnancy and the Postpartum Period: A Multivariate Bayesian Meta-Analysis. J Clin Psychiatry. 2019 Jul 23;80(4):18r12527.

66. Dennis CL, Falah-Hassani K, Shiri R. Prevalence of antenatal and postnatal anxiety: systematic review and meta-analysis. Br J Psychiatry J Ment Sci. 2017 May;210(5):315–23.

67. Waring ME, Bhusal S, McManus-Shipp KE, Field CM. Engagement with digital psychoeducational content to promote mental health and well-being among perinatal persons and mothers [dataset] [Internet]. Ann Arbor, MI: Inter-university Consortium for Political and Social Research; 2024. Available from: https://www.openicpsr.org/openicpsr/project/204321

